# Network analysis of the whole body kinematics in Parkinson diseases following levodopa intake

**DOI:** 10.1101/2023.06.29.23292021

**Authors:** Antonella Romano, Marianna Liparoti, Roberta Minino, Arianna Polverino, Lorenzo Cipriano, Francesco Ciaramella, Anna Carotenuto, Domenico Tafuri, Giuseppe Sorrentino, Pierpaolo Sorrentino, Emahnuel Troisi Lopez

## Abstract

**Background:** Three-dimensional motion analysis represents a quantitative and objective approach to assess spatio-temporal and kinematic changes in Parkinson’s disease (PD). However, these parameters, focusing on a specific body segment, provide only segmental information, discarding the complex whole-body patterns underlying the motor impairment. We aimed to assess how levodopa intake affects the whole body, large-scale kinematic network in PD.

**Methods:** Borrowing from network theory, we used the kinectome framework by calculating the Pearson’s correlation coefficients between the time series acceleration of 21 bone markers. Then, we performed a topological analysis to evaluate the large-scale interactions between body elements. Finally, we performed a multilinear regression analysis in order to verify whether the kinectome’s topological features could predict the clinical variation before and after levodopa intake.

**Results:** PD patients showed lower nodal strength (i.e., lower synchronization) in the upper body in the medio-lateral acceleration while in on-state with respect to the off state (p-head=0.048; p-C7=0.032; p-T10=0.006). On the contrary, PD patients in on state displayed higher nodal strength (i.e., higher synchronization) of both elbows (right, p=0.002; left, p=0.005), wrists (right, p=0.003; left, p=0.002) and knees (right, p=0.003; left, p=0.039) in the antero-posterior acceleration. Furthermore, the predictive analysis revealed that the nodal strength variations of the arms, following levodopa intake, significantly predicted the clinical variations assessed through the UPDRS-III (R2=0.65; p=0.025).

**Conclusions:** PD patients in the on-state showed less rigidity during walking, proportional to the UPDRS-III variation. More importantly, we showed that levodopa induces an improvement of the whole body, large-scale kinematic pattern.

## Introduction

Parkinson’s disease (PD) is neuropathologically characterized by the loss of dopaminergic nigro-striatal neurons ^1^. However, PD is now recognized as a multi-system, multi-neurotransmitter dysfunction-related heterogeneous disorder, and this heterogeneity may probably be mirrored by the different observable clinical phenotypes, culminating in a complex motor and nonmotor disorder ^2^.

Motor impairment, which remains predominant in the clinical picture and drastically affects the quality of life ^3^, is characterized by several cardinal symptoms, including reduced balance and hampered coordination which often leads to falls ^4^

To this regard, the assessment of voluntary movement is crucial to achieve early diagnosis, to estimate motor disability, and to track disease progression, as well as the effects of rehabilitation and pharmacological interventions. However, the motor evaluation in PD predominantly relies on the individual expertise of the clinician. To overcome this limitation, the Unified Parkison’s disease Rating Scale (UPDRS, part III) is widely used. ^5^. However, similarly to the clinical evaluation, the UPDRS suffers from several, indisputable, limitations, which, once again, are related to the subjectivity of the investigator.

Nowadays, the three-dimensional motion analysis (3D-MA), by investigating spatiotemporal, kinetic and kinematic parameters ^6–8^, is considered the gold standard for fine-tuned motor assessment in PD (as well as other neurological diseases) and, in particular for gait alterations. For example, it has been shown that PD patients exhibited a reduced step length and step speed as well as an increased double-limb support time with respect to healthy controls and, from a kinematic standpoint, a reduction of the ankle joint angular excursion ^9^. An alternative approach has led to the development of synthetic biomechanical indices that enable the analysis of more complex characteristics of gait such as the fluidity, the rhythmicity and the symmetry ^10–13^. However, the majority of these methodological approaches provide only “segmental” information, by selectively focusing only on a specific body segment such as the trunk or legs. In other words, the major constraint of these approaches is that they provide synthetic final outcomes that do not take into account the complex patterns that generated the movement itself, thus leading to a loss of information. Therefore, an accurate characterization of movement patterns alterations in PD requires not only precise measurements, but also appropriate mathematical methods able to conceptualize the movement of the human body as a complex system whose components are highly interconnected to each other ^14^.

Network theory is a solid, methodological framework able to describe the relationship between the elements of a complex system defining not only its properties as a whole, but also the contribution of each element within the network itself ^15^. Hence, network theory could be a suitable approach able to describe the whole body interactions underlying the motor patterns in health and motor disease, such as PD. To this regard, the *kinectome* framework has been recently developed in order to provide a comprehensive, large-scale description of human gait kinematics through the analysis of the complex interactions between the body segments that generated the movement patterns. (Troisi Lopez et al., 2022). The kinectome stems from the application of network theory to human movement ^17^ and allows the investigation of the kinematic interactions occurring between anatomical segments during movement.

In PD, the kinectome analysis revealed that there is a greater dysregulation in the whole-body movement pattern (i.e., higher variability) during gait with respect to healthy controls. That is, at the whole body level, PD causes an alteration of the synergies among anatomical body parts which, more importantly, is correlated with the clinical evolution of disease assessed through the UPDRS-III (Troisi Lopez et al., 2022).

Although the pioneering studies by Hornykiewicz and Birkmayer ^18^ on the use of levodopa in the treatment of PD date back to the early 1960s, levodopa remains the gold standard in therapeutic management (Jankovic, 2008) and its efficacy in improving motor symptomatology is one of the main diagnostic criteria. For example, the assumption of levodopa results in a reduction of the variability coefficient of several spatiotemporal parameters (i.e., the stride length and the swing velocity) ^19,20^. However, while the efficacy of levodopa in relieving specific aspects of motor impairment, such as bradykinesia, is widely established, the drug’s effects on whole body kinematics are poorly studied, resulting in a lack of information on the motor aspects that mostly impair the quality of life (i. e., balance reduction and hampered coordination). To overcome such limitations, Troisi Lopez et al., developed a synthetic kinematics index of trunk displacement as a measure of postural stability, named TDI which measures the displacement of the trunk with respect to the center of mass (CoM). Nevertheless, the TDI is still not able to evaluate large scale, whole body dynamic balance alterations during gait. (Troisi Lopez et al., 2021).

To our knowledge, a comprehensive analysis of more complex gait features, such as turning and subtle balance adjustment during gait, is still lacking ^21^, similarly to a whole body network analysis assessing changes in the kinematic network following the assumption of pharmacological therapy.

To this aim, we used a 3D-MA system and reflective markers, to obtain the acceleration time series of several body segments of twenty-three PD patients who were recorded before and after the intake of a subclinical dose (half of the morning dose) of levodopa. Hence, we built the kinectome by calculating the covariance between each pair of bone markers. Then, we performed a topological analysis to explore a possible variation of the large-scale interactions among body elements due to the assumption of the antiparkinsonian treatment. Finally, we performed a multilinear regression analysis to check whether these topological variations were related to the clinical variations assessed through the UPDRS-III.

## Materials and Methods

### Participants

Twenty-three patients affected by Parkinson’s disease (17 males and 6 females; mean age 65.3±11.58; education level 10.73±3.84) (Table 1) were recruited from the Movement Disorder Unit of the Cardarelli Hospital in Naples. The PD diagnosis was defined according to the United Kingdom Parkinson’s Disease Brain Bank criteria ^22^. Inclusion criteria were: (1) minimum age of 45 years or older; (2) Hoehn and Yahr (H&Y) ^23^ score ≤3 while at off state (i.e., without any antiparkinsonian treatment); (3) disease duration <10 years; (4) antiparkinsonian treatment at a stable dosage (5) absence of any neurological (except for PD) or psychiatric disorder. Exclusion criteria were: (1) Mini-Mental State Examination (MMSE) < 24 ^24^; (2) Frontal Assessment Battery (FAB) <12 ^25^; (3) Beck Depression Inventory II (BDI-II) >13 ^26^; (4) assumption of psychoactive drugs; (5) any physical or medical conditions causing walking impairment. The study protocol was approved by the local ethics committee “Azienda Ospedaliera di Rilievo Nazionale A. Cardarelli” (protocol number: 0001962) and all participants provided written informed consent in accordance with the Declaration of Helsinki.

**Table 1:**
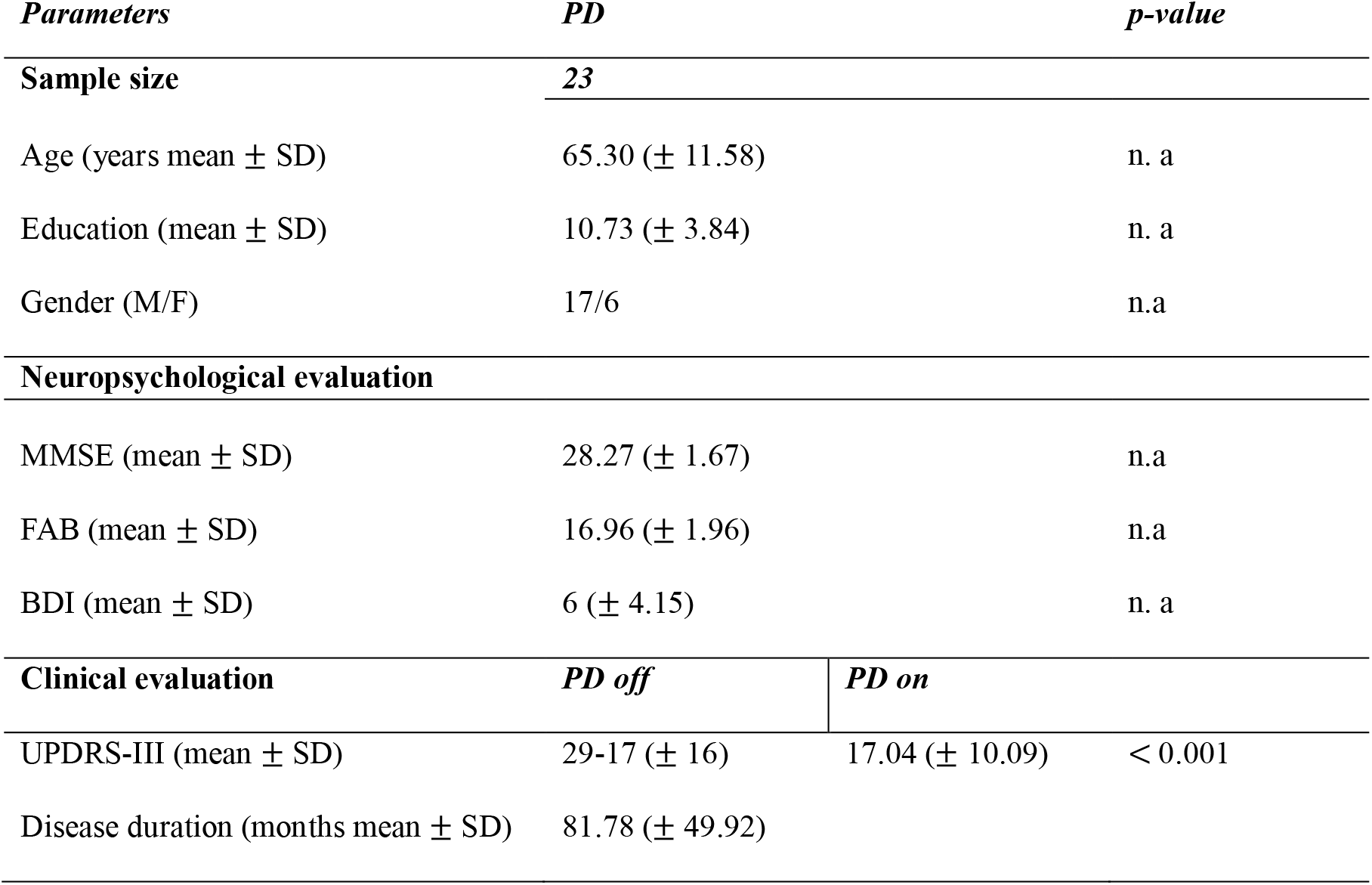
Participants characteristics. MMSE: Mini Mental State Examination; FAB: Frontal Assessment Battery; BDI: Beck Depression Inventory; UPDRS-III: Unified Parkinson’s disease rating scale part three. n.a: not available

### Stereophotogrammetric acquisition

The acquisitions took place in the Motion Analysis Laboratory (MoveNet Lab) of the University of Naples Parthenope. Gait data were obtained through a stereophotogrammetric system composed by eight infrared cameras (ProReflex Unit—Qualisys Inc., Gothenburg, Sweden) with a sampling frequency of 120 frames per second. Fifty-five passive markers were positioned on the naked skin of participants in specific anatomical landmarks according to the modified Davis protocol ^27^. We asked the participants to walk straight at their preferred speed through a measured space (10 meters). PD patients were recorded twice: during the first acquisition, PD patients were in off state (i.e., no antiparkinsonian treatment in the last 14-16 hours). The second acquisition was performed 40 minutes after patients had taken a subclinical dose (i.e., half of their usual morning intake) of levodopa (Malevodopa+Carbidopa) (on state). Before each acquisition each participant wes tested through UPDRS-III ^28^. For both conditions (i.e., off state and on state) we recorded four gait cycles including two complete right and left gait cycles. Hence, we obtained the acceleration time series for each bone marker.

### Kinectome analysis

Here, we applied the recently developed *Kinectome* framework ^16^ to provide a comprehensive description of the large-scale gait features in Parkinson’s disease and to investigate how the levodopa intake affects the large-scale movements in Parkinsonians. Overall, for each patient, we obtained six kinectomes (2 conditions x 3 axes), using the acceleration time series alongside the three axes of movement (i.e., vertical (VT), anteroposterior (AP) and mediolateral (ML). The Kinectome is a covariance matrix obtained by calculating the Pearson’s correlation coefficients between each couple of time series, among 21 bone markers. Hence, we identified the bone markers as nodes while the links (i.e., the edges) were defined by the level of synchronization between each couple of nodes (i.e., the corresponding correlation coefficient). Thereby, we obtained a symmetric matrix containing 420 edges (Figure 1), where the elements on the diagonal are equal to 1 since they represent the correlation of a node with itself.

**Figure 1:**
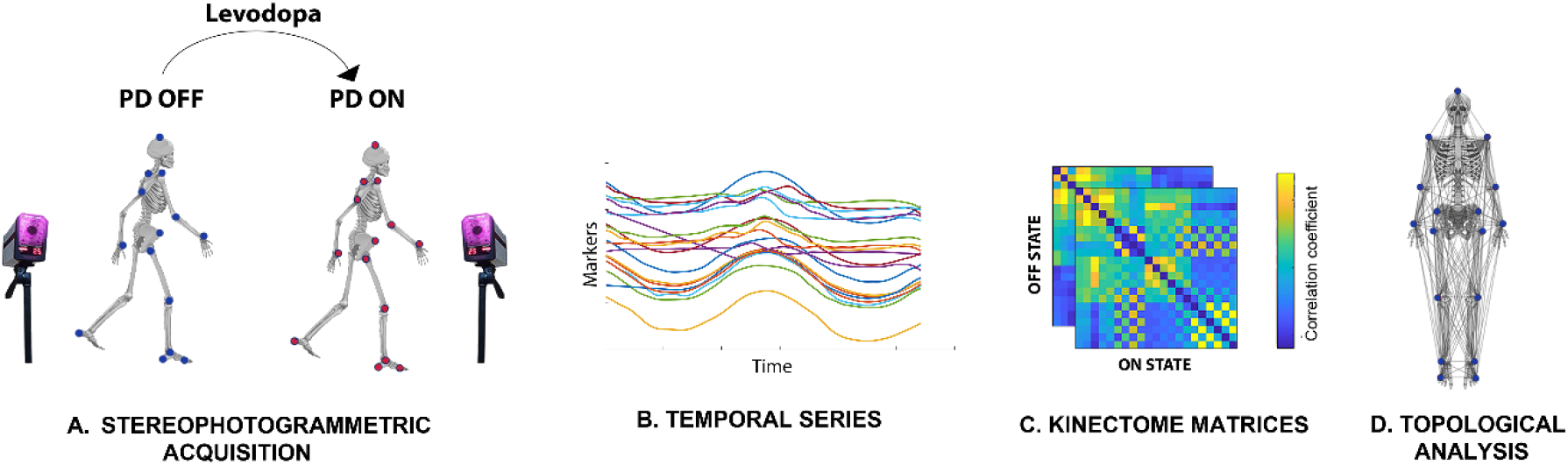
Pipeline overview. **A. Stereophotogrammetric acquisition**. 23 PD patients were recorded through a stereophotogrammetric system before (PD off) and after (PD on) the assumption of levodopa. Blue and red dots represent the anatomical position of the bone marker. **B. Temporal series**. The time series of the acceleration are obtained from the bone markers’ position during the gait cycle. **C. Kinectome matrices**. For each patient we obtained two kinectomes (i.e., ON state and OFF state) alongside the three axes of movement (VT, AP, ML) by computing the correlation coefficient between each pair of time series. **D. Topological analysis**. Bone markers network representation used for the topological analysis.

### Topological analysis

Through the kinectome we wanted to conceptualise the whole body as a network in which all body parts are mutually dependent. Hence, borrowing from graph theory and network analysis, we performed a topological analysis in order to assess the role of each anatomical bone marker with respect to the whole body (i.e., the synchronization level of a body element with respect to the other ones). Specifically, we calculated the nodal strength which represents the topological importance of a given node within the kinematic network ^29^. The nodal strength is calculated as:

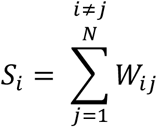

where *i* and *j* represent two nodes in the network, *W* is the edge connecting them and *N* is the total number of the nodes of the network.

### Multilinear regression analysis

We used the topological features obtained from the kinectome analysis in order to predict the clinical variations induced by the levodopa intake. To this end, we built a multilinear regression model in which the UPDRS-III variations (i.e., Δ-UPDRS-III) (UPDRS-III in off condition - UPDRS-III in on condition) represented the dependent variable, while age, gender, education level, disease duration (expressed in months) and the topological features of interest were the predictors. Multicollinearity was assessed through the variance inflation factor (VIF). To validate our approach, we performed *k*-fold cross-validation, with *k* = 5 ^30^. Specifically, *k* iterations were performed to train our model and at each iteration the *k*^*th*^ subgroup was used as a test set. The cross-validation procedure was repeated one hundred times to exclude that the result was caused by random sampling.

### Statistical analysis

Statistical analysis was carried out in Matlab (Mathworks version 2021a). A two-sided Wilcoxon signed rank test was performed to compare the topological features between the two conditions (i.e., PD on state and PD off state). The results were corrected for multiple comparisons using the False Discovery Rate (FDR) method ^31^. Significance level was set at p < 0.05.

## Results

### Nodal strength investigation

We performed a topological analysis in order to verify whether the levodopa intake in PD patients resulted in a change of the synchronization level of a given node (i.e., a bone marker) with respect to the other body segments. We found statistical differences in ML and AP axes. Concerning the ML axis, significant differences were present in the upper part of the body, showing greater synchronization in PD while in off condition. Conversely, significant results in the AP axis showed greater synchronization in on condition and involved both upper and lower limbs. These results highlight a reduction of the ML hyper-synchronization of the trunk (i.e., rigid oscillations of the upper body), in favour of a better coordination of upper and lower limbs on the AP axis (Figure 2).

**Figure 2.**
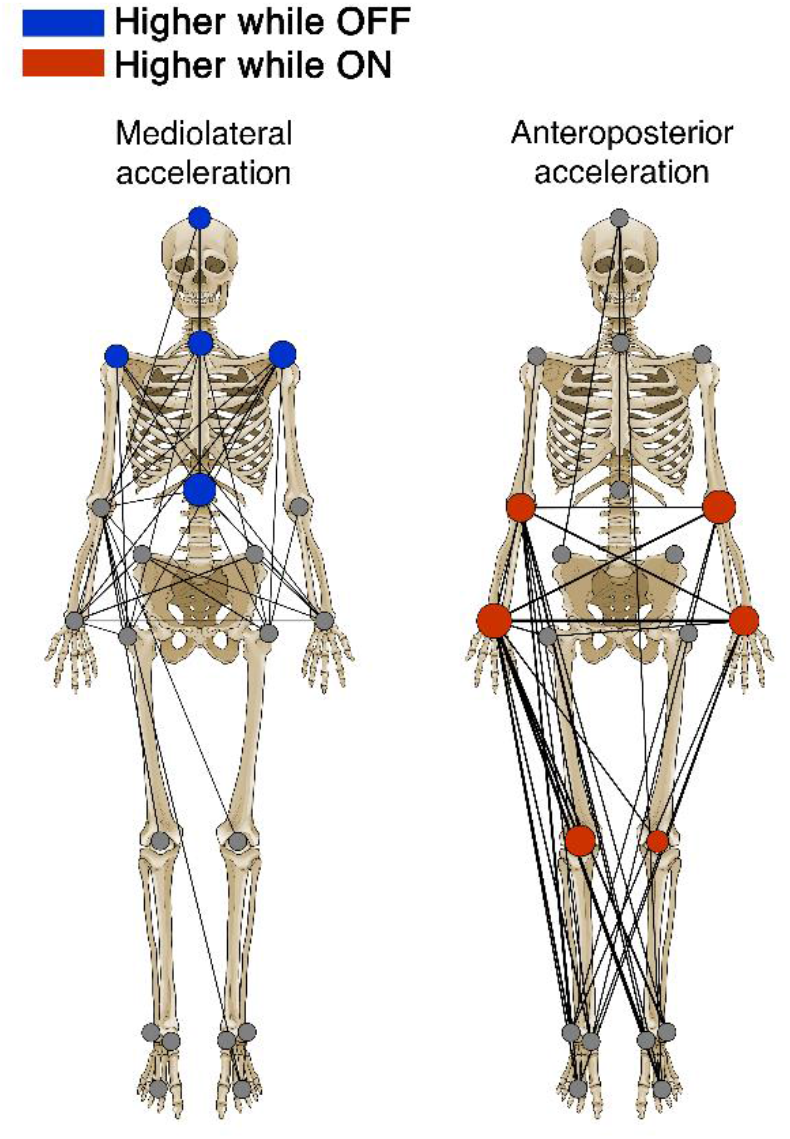
Visual representation of the statistical differences. The two skeletons highlight the significant differences of the topological feature between off and on conditions. The left skeleton displays differences in the mediolateral axis, while the right skeleton displays differences in the anteroposterior axis. Colored nodes represent significant differences between off and on conditions (i.e., without and with levodopa medicament, respectively) in patients with Parkinson’s disease. Blue nodes whether synchronization was higher in off condition, red nodes whether synchronization was higher in on condition. The size of the colored nodes depends on the effect size (z value) of the statistical comparison. Gray nodes represent no significant difference. Furthermore, 25% of the network’s links with the higher values are displayed. Thickness of the link depends on the respective edge value.

Specifically, on the ML axis, after the levodopa intake, the patients exhibited reduced nodal strength values with regard to the head (pFDR = 0.048), the 7^th^ cervical vertebra (C7) (pFDR = 0.032), the 10^th^ thoracic vertebra (T10) (pFDR = 0.006) and the right (RAC) and the left acromion (LAC) respectively (pFDR = 0.040; pFDR = 0.027) (Figure 3). On the contrary, after levodopa intake, on the anteroposterior axis, the PD patients exhibited higher nodal strength values of both the right (RLELB) and left (LLELB) elbows (pFDR = 0.002; pFDR = 0.005), the right (RWRB) and left (LWRB) wrist (pFDR = 0.003; pFDR = 0.002) and the left (LLK) and right lateral knee (RLK) (pFDR = 0.039; pFDR = 0.003) (Figure 4). Therefore, the levodopa intake resulted in a reduction of the synchronization level of the body segments at the trunk level (i.e., lower nodal strength values) with respect to the rest of the body, and, conversely, an increase of the synchronization level (i.e., higher nodal strength values) of both the right and the left arm and the right and left knee respectively.

**Figure 3:**
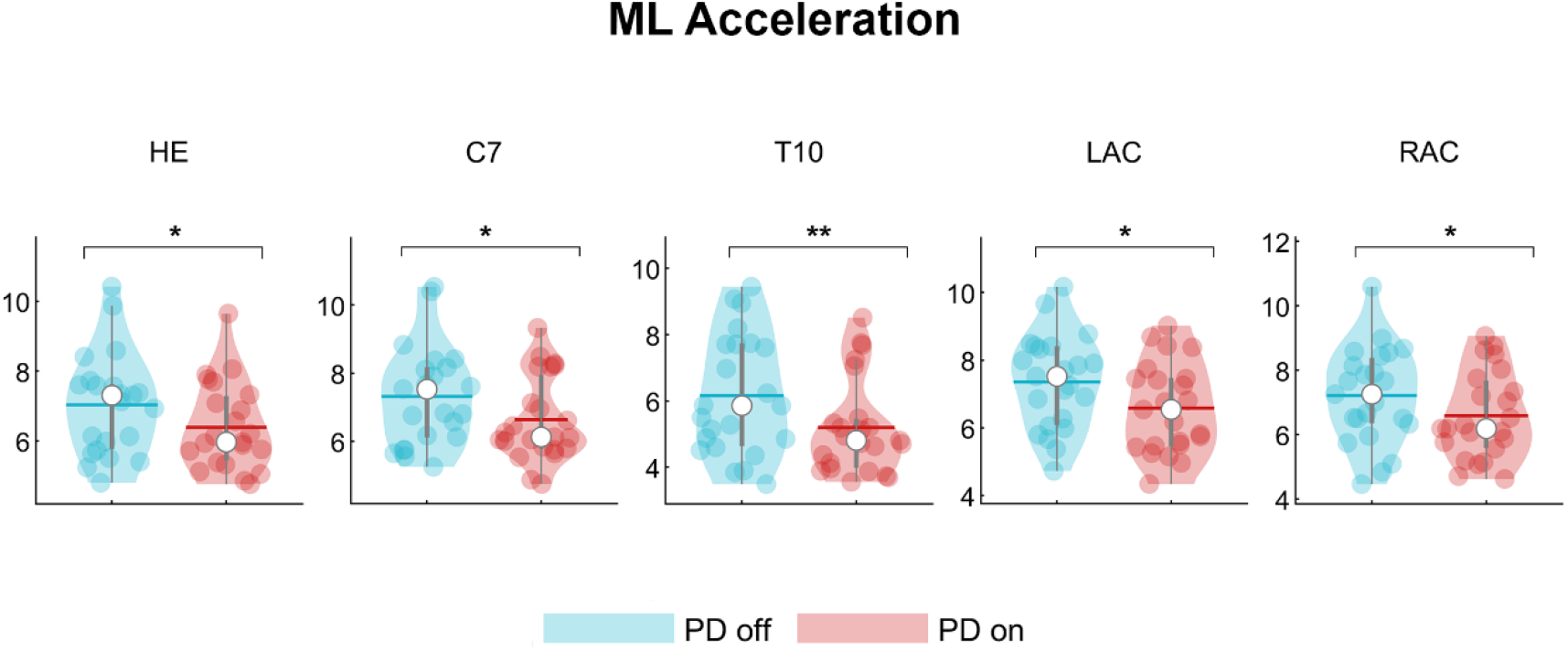
Topological comparison in mediolateral acceleration. Violin plots represent the nodal strength comparison between PD patients in off state and PD patients in on state. PD patients during the off state showed higher values of nodal strength of the head (HE), the 7^th^ cervical vertebra (C7), the 10^th^ thoracic vertebra (T10) and the left (LAC) and the right (RAC) acromion with respect to the PD patients in the on state. *p < 0.05; **p < 0.01.

**Figure 4:**
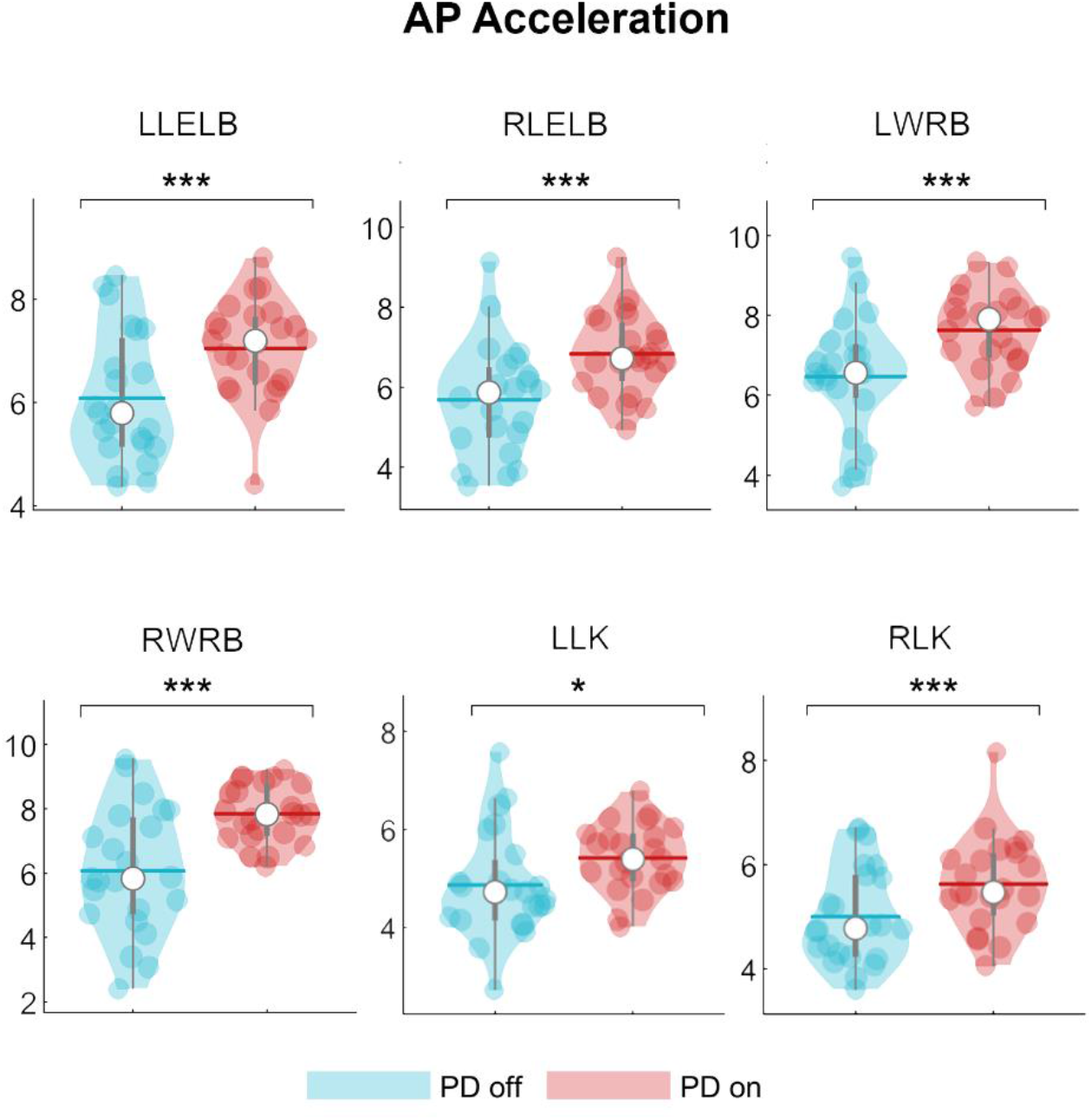
Topological comparison in antero-posterior acceleration. Violin plots represent the nodal strength comparison between PD patients in off state and PD patients in on state. Following the levodopa intake, PD patients showed higher nodal strength values of the left (LLELB) and right (RLELB) elbows, the left and right wrists (LWRB, RWRB) and left and right knees (LLK, RLK) with respect to the PD patients in the on state. *p < 0.05; ***p < 0.001.

### Clinical prediction

Based on previous findings ^16^, we aimed to test if the clinical variation (Δ-UPDRS-III) could be related to the variation of the ML-T10 nodal strength (Δ-T10). To this end, we built a multilinear regression model, validated through k-fold cross validation (k = 5) in which Δ-T10 was the predictive variabile (among other nuisance predictors such as age, education level, gender, disease duration) and the Δ-UPDRS-III was the responsive variable. The Δ-T10 did not predict the Δ-UPDRS-III (p = 0.332; β = 0.320; R2 = 0.17) (data not shown).

However, as we have previously shown, the levodopa intake resulted in a change in the motor pattern of the PD patients characterised by a decrease of the synchronization at the trunk level and an increase of the synchronization level of the upper limbs in the AP axis. Hence, we wondered whether the clinical changes (Δ-UPDRS-III) could be mirrored by the AP variations of the upper limbs’ nodal strength values (i.e., Δ-RLELB; Δ-LLELB; Δ-RWRB; Δ-LWRB). Our result showed that the nodal strength variations of the right elbows and both the left and right wrist significantly contributed to the prediction of the Δ-UPDRS (RLELB p = 0.002, β = -1.052; LLELB p = 0.009, β = -0.768; RWRB p = 0.01, β = 0.888; R2= 0.65) (figure 5).

**Figure 5:**
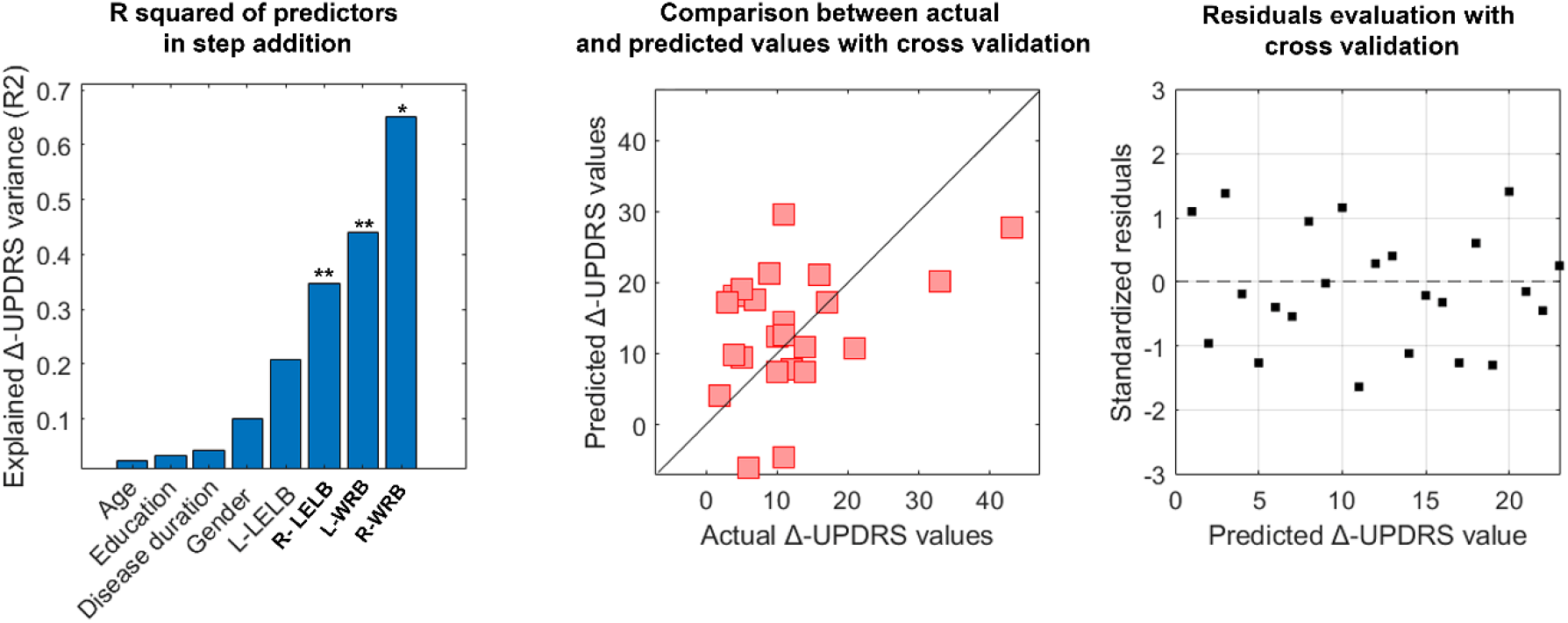
Clinical prediction. Multilinear regression analysis with *k-fold* cross validation was performed to verify the ability of the nodal strength of the upper limb to predict the clinical variation of the UPDRS-III before and after the levodopa intake (i.e., PD off - PD on). The left column displays the explained variance obtained by adding the predictors (age, education level, disease duration, gender and the nodal strength values of the right and left elbow and the right and left wrist). The central column displays the comparison between the predicted and the actual values of the responsive variable validated through the *k-*fold cross validation. Finally, in the right column, is displayed the distribution of residuals representing the standardisation of the difference between the actual and predicted Δ-UPDRS-III values. The significant predictors are highlighted in bold while the significant *p*-value is indicated with * (p < 0.05), **(p< 0.01).

## Discussion

In the present study, we used the recently developed kinectome framework to explore how the assumption of levodopa affects the large-scale kinematic interactions between body segments during gait in PD. Firstly, starting from the temporal series of the acceleration, we obtained, for each patient, the covariance matrices (i.e., the kinectomes) which estimated the level of synchronization between pairs of body segments (through Pearson’s correlation). In the second place, we performed a topological analysis in order to investigate the role of each human kinematic element with respect to the other ones during gait and its possible variations after the assumption of levodopa. Finally we aimed to verify whether these topological variations could be related to the clinical variation assessed through the UPDRS part III.

Our results revealed that, while in the on-state, PD patients showed a reduction of the nodal strength (i.e, lower synchronization) of the trunk (i.e., head, acromions and back) in the ML-A and, conversely, an increase of the nodal strength (i.,e higher synchronization) of the upper limbs (wrists and elbows) and the knees in the AP-A with respect to the off state. Hence, following the pharmacological treatment, PD patients exhibited a lower coordination of the trunk with respect to the whole body and a greater coordination between upper and lower limbs during walking. Our results are coherent with our previous findings. Indeed in Trosi Lopez et al., we found that PD patients in off state, showed an hypersynchronization of the trunk (i.e., increased nodal strength of the 10th thoracic vertebrae) representative of the rigidity in PD as confirmed by the positive correlation with the UPDRS-III. Here we observed that, following the assumption of levodopa, the trunk becomes more independent (i.e., less synchronized) with respect to the other body segments, showing the ability of levodopa to improve the rigidity and the smoothness of gait ^1,16,20,21^.

Inter-limbic coordination, which is essential to provide dynamical stability and smoothness during gait ^32^, is disrupted in patients with PD ^33^. Indeed it has been shown that both ipsilateral and contralateral coordination was altered in PD and appeared to be related to a worse clinical condition (assessed through the UPDRS-III) ^34^. Moreover, Winogrodzka and colleagues showed that PD patients with enhanced inter limbs coordination deficits were those in a more advanced stage of disease which displayed greater bradykinesia and rigidity in contrast with early or drug-naive PD patients who, through the manipulation of gait speed, showed a better preservation of the inter-limbic coordination ^35^. Our results are in line with a previous study by Son et al. who showed that the assumption of levodopa led to an increase of the phase coordination index which was also related to a PD patients’ better clinical condition and a greater stability ^36^.

Previous studies have demonstrated that levodopa treatment can reduce variability in PD by acting on the dopaminergic pathways ^21,37,38^. For instance, Park et. al., suggested that the nigrostriatal dopaminergic modulation could play a central role in the formation of locomotor synergies (i.e., a neural organization), which are responsible for the development of movement pattern and stability ^39,40^. Indeed, in another study by Carpinella et al., it has been shown that Subthalamic Nucleus Stimulation alongside the levodopa assumption led to an improvement of coordination between upper and lower limbs ^41^.

Hence, we can speculate that the improvement of motor pattern synchronization may be due to the ability of the pharmacological treatment to supply to the impairment of those brain areas involved in the synchronization and sequencing of movements such as the Basal Ganglia and/or the Supplementary Motor Area ^42–45^. Further investigations are needed to deepen the actual pharmacological effects of levodopa on cortical areas involved in movement coordination.

Finally, we performed two multilinear regression analyses to check any clinical relationship between the topological variations assessed through the kinectome analysis and the UPDRS-III. The first predictive model revealed that the Δ-T10 nodal strength was not predictive of the UPDRS-III variation (i.e., UPDRS-III off - UPDRS-III on). We used the T10 nodal strength as predictor due to its strong relationship with the clinical assessment of the disease in the off state ^16^. It is quite expected that the variation T10 nodal strength is not predictive of the Δ-UPDRS-III since, when PD patients turn from the off to the on state, the nodal strength values decrease in a way that is not directly linked to the clinical motor score but that depends on several factor such as the subject-specific response to levodopa. To this regard, it is important to highlight that the nodal strength value of a bone marker is representative of the weight (i.e., the importance) that a node plays in the harmony and fluidity of a movement pattern compared to the rest of the body segments. This would explain why the nodal strength variation of T10 does not carry any predictive power.

The second multilinear regression model showed that, among other nuisance predictors, the nodal strength variations of the arms (i.e., left and right wrist and right elbow) was significantly able to predict the ΔUPDRS-III. This means that PD patients in the on-state showed less rigidity during walking which was proportional to the UPDRS-III variation. Hence, the AP acceleration movements of the arms play a fundamental role in enhancing smoothness in patients with PD.

A converging line of evidence assesses the role of the upper limbs in walking in both health and disease. Indeed, arms swing is essential to minimize the energy expenditure as well as to improve dynamic stability ^46–49^. Intriguingly, it has been shown that upper limb movement influences the recruitment of lower limbs during rhythmic activities (e.g., walking) ^47^. Please note that arm swing symmetry and coordination is disrupted in PD ^50,51^. However, in their study, Warmerdam et al., showed that, following the levodopa assumption, PD patients exhibited an improved arm swing, especially for what concerned the main amplitude, the peak angular velocity, coordination and sideway amplitude ^52^, suggesting, in agreement with our results, that, following pharmacological treatment, the arm swing may occur to facilitate gait pattern in PD patients.

## Conclusion

In the present work we used the recently developed kinectome framework to explore large-scale kinematic changes in patients with PD following the assumption of levodopa. Our results revealed that, at whole body level, the levodopa intake led to an enhanced synchronization between the upper and lower limbs which was predictive of the UPDRS-III variation. We hope that this approach may be helpful in monitoring subtle, whole body changes during walking in PD patients due to the pharmacological treatment and, more importantly, that future rehabilitative approaches may focus on the upper limbs due to the role that they play in the fluidity of motor gestures.

## Data Availability

All data produced in the present study are available upon reasonable request to the authors

## Acknowledgments

We would like to thank Bronwen Hughes for the english revision of the manuscript.

## Authors contribution

Antonella Romano and Marianna Liparoti: Conceptualisation, data acquisition, data processing, writing-original draft preparation; Lorenzo Cipriano, Roberta Minino, Arianna Polverino, Francesco Ciaramella: data acquisition, results interpretation, manuscript revision; Anna Carotenuto: enrolment; Domenico Tafuri: results interpretation and manuscript revision; Giuseppe Sorrentino: study conceptualisation, supervision and funding; Pierpaolo Sorrentino: Conceptualisation, writing-original draft preparation, study supervision and funding; Emahnuel Troisi Lopez: Conceptualisation, data processing, writing-original draft preparation and supervision.

